# COVID-19 in regions with low prevalence and low density of population. An uncertainty dynamic modeling approach

**DOI:** 10.1101/2020.10.21.20215418

**Authors:** José M. Garrido, David Martínez-Rodríguez, Fernando Rodríguez-Serrano, Javier Díez-Domingo, COVID-19_Granada Study Group, Rafael-J. Villanueva

## Abstract

The coronavirus disease 2019 (COVID-19) that emerged in China at the end of 2019 has spread worldwide. In this article, we present a mathematical SEIR model focused on analysing the transmission dynamics of COVID-19, the patients circulating in the hospitals and evaluating the effects of health policies and vaccination on the control of the pandemic. We tested the model using registered cases and population data from the province of Granada (Spain), that represents a population size near 1 million citizens with low density of population and low prevalence. After calibrating the model with the data obtained from 15 March to 22 September 2020, we simulate different vaccination scenarios - including effectiveness and availability date - in order to study the possible evolution of the disease. The results show that: 1) infected will increase until 5.6% - 7.4% of the total population over next 3-4 months (2nd wave); 2) vaccination seems not to be enough to face the pandemic and other strategies should be used; 3) we also support the claim of the WHO about the effectiveness of the vaccine, that should be, at least, of 50% to represent a substantial progress against the COVID-19; 4) after the 2nd wave, the return to normal life should be controlled and gradual to avoid a 3rd wave. The proposed study may be a useful tool for giving insight into the transmission dynamics of SARS-CoV-2 and to design vaccination and health policies.

## 1 Introduction

Last December, the Chinese public health authorities informed about several cases of acute respiratory syndrome in Wuhan. By 7 Jan 2020, scientists had isolated a novel coronavirus from these patients not previously identified in humans, and referred as severe acute respiratory syndrome coronavirus 2 (SARS-CoV-2), the agent responsible for the later designated coronavirus disease 2019 (COVID-19) in February 2020, and the World Health Organization (WHO) declared a pandemic [1, 2]. Much is known about the virus and the disease, but there is a crucial need to further knowledge about the virus transmission to control the spread and to implement measures to reduce the impact, like social distancing or the use of facemasks, while effective antivirals and vaccines are obtained.

Different mathematical models have been proposed to analyse the evolution of this pandemic although with limitations due to the many uncertainties regarding to the disease. One of the main parameters in the epidemiological models is the effective reproductive number (*R*_0_), which quantifies the average of secondary cases generated from each infected person in a susceptible environment during an outbreak. Different studies have estimated that the *R*_0_ value in China at the beginning of the pandemic was between 2 and 7.1, and also it has been observed that *R*_0_ value drops sharply after the implementation of control measures against COVID-19 [3].

Several approaches have tried to model mathematically the transmission dynamics of COVID-19 [4, 5, 6, 7]. Other references as [8, 9, 10, 11], moreover, consider asymptomatic infected individuals, most of the infected population, who play an important role in spreading the virus.

In order to make comparable our study with other similar regions, we are going to give our results in absolute values and percentages. Also, under a demographic point of view, Granada is a province in southern Spain with a population of 921511 inhabitants, around 30% of the population live in the capital and the province has a population density of 73.5*/km*^2^ [12] Compared with the province of Madrid with 829.6*/km*^2^, the province of Granada has a low density of population. And the percentage of accumulated infected between 1 May 2020 and 1 June 2020 was [2.3% - 4.8%] in Granada and [10.3% - 13.3%] in Madrid [13]. Therefore, the province of Granada has a low prevalence.

In this paper we propose a mathematical model to study the current transmission dynamics of SARS-CoV-2. It is a SEIR model in which the circuit of patients moving throughout the hospital dependencies is also considered, returning a more precise portrait of the use of hospital resources with the aim of predicting hospital’s overloads. This is a novelty that our model provides and any other of the models in the literature, to our knowledge, do. Also, we consider the asymptomatic infectious individuals who transmit the disease but not reported by the system. We proved the model using registered hospital’s cases and population data from the province of Granada (Spain). Once the model was implemented and calibrated, we performed calculations simulating possible future scenarios to evaluate the effect of the COVID-19 vaccination, that allow us to provide some public health recommendations.

## 2 Model building

With respect to COVID-19, an individual can be considered as:

- **(S)** susceptible, when the individual is healthy;
- **(Q)** in lockdown, when the individual is at home to avoid the spread of the virus. In our model, lockdown only considers the lockdown time;
- **(L)** latent or exposed, when the individual has been infected but it is not infectious yet; infectious, when the individual is capable of spreading SARS-CoV-2;
- **(R)** recovered, when the individual recovers from the disease being asymptomatic or having mild symptoms;
- **(H)** hospitalized at ward, when the individual has severe symptoms and needs to be hospitalized;
- **(u)** in intensive care unit (ICU), when the individual has severe symptoms and needs to be treated in the intensive care unit;
- **(F)** deceased, when the individual dies because of the disease;
- **(HU)** after ICU, when an individual is transferred from ICU to other non-ICU department due to improvement in the evolution but still requires hospitalization;
- **(A)** discharged, when the individual gets better and is discharged from hospital.

We are going to use data of the Spanish province of Granada provided by the hospitals H.U. Virgen de las Nieves, H.U. Clínico San Cecilio, H.U. Santa Ana in Motril, H.U. of Baza, H. of San Rafael and H.L.A. Inmaculada, generating an inclusive model which represents the whole province. The province of Granada has *P*_*T*_ = 921 511 inhabitants, and we consider the *P*_*T*_ value as constant during the simulation time. Our simulations begin on 1 March 2020 and the time step *t* is one day.

1. When the alarm state is decreed and most of the people have to be in lockdown, that is, move from *S* to *Q*, it is modeled by the term *s*_*q*_(*t*), where the model parameter *s*_*q*_(*t*) determines the transit of people from *S* to *Q. s*_*q*_(*t*) takes the value 0 except for 16 March 2020 and 31 March 2020, when the lockdown and the strict lockdown began in Spain, and 700 000 and 150 000 individuals in Granada, respectively, move from S to Q [14].
2. When the lockdown finishes, the transit of individuals from *Q* to *S* is modeled by the term *q*_*s*_(*t*) where the model parameter *q*_*s*_(*t*) is 0 except for 13 April 2020 when the strict lockdown finishes and 150 000 individuals move from Q to S, and from 5 May to 21 June 2020, leaving the lockdown 8 750 people every day due to the gradual end of the confinement.
3. An individual moves to latent state (L) if he/she gets infected by contact with an infectious contagions. The transit is modeled by the non-linear term 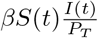, where the transmission rate parameter *β* has to be calibrated. Furthermore, this parameter will change over time due to the global public health interventions.
4. A latent individual transits to infectious state after a while and this is modeled by the linear term *l*_*i*_*L*(*t*), where the latency period *t*_0_ = 1*/l*_*i*_ is the time from the moment in which an individual is infected until the moment in which is able to transmit the virus, *t*_0_. This period is different to the typical incubation period time from infection to onset, and according to [15], it takes from 2 to 7 days. Even though there is limited evidence about the possibility of infection one or two days before onset [16], let us consider that *t*_0_ may take values from 1 to 6 days, and *l*_*i*_ = 1*/t*_0_.
5. An infectious individual may become hospitalized (H), admitted to Intensive Care Unit (U) or get recovered (R), and these transits are modeled by the linear terms *i*_*h*_*I*(*t*), *i*_*u*_*I*(*t*) and *i*_*r*_*I*(*t*), respectively. Here we have 3 possible ways for infectious individuals. Every one takes its time and has its probability, that is, *i*_*h*_ = *p*_1_*/t*_1_, *i*_*u*_ = *p*_2_*/t*_2_ and *i*_*r*_ = (1 *− p*_1_ *− p*_2_)*/t*_3_.
  - *p*_1_ is the percentage of infected who become hospitalized and *p*_2_ is the percentage of infected people admitted directly at ICU. Then, the parameters of the transition from *I* to *H* and from *I* to *U* are *i*_*h*_ = *p*_1_*/t*_1_ and *i*_*u*_ = *p*_2_*/t*_2_, respectively.
  - Those infected not hospitalized require around *t*_3_ = 14 days to recover [17]. Thus, the transition from *I* to *R* is governed by the parameter *i*_*r*_ = (1 *− p*_1_ *− p*_2_)*/*14.
6. People hospitalized may move to ICU (U) if get worse, may decease (F) or may be discharged (A). These transits are modeled by the linear terms *h*_*u*_*H*(*t*), *h*_*f*_ *H*(*t*) and *h*_*a*_*H*(*t*), respectively. As before, here we have 3 possible ways for hospitalized individuals.
  - *p*_4_ is the percentage of hospitalized people who need to be admitted to ICU. Hence, the people in *H* move to *U* governed by the parameter *h*_*u*_ = *p*_4_*/t*_4_.
  - The transition parameter from *H* to *F* is *h*_*f*_ = *p*_5_*/t*_5_.
  - (1 *− p*_4_ *− p*_5_) of hospitalized are discharged after an average of *t*_6_ days in the hospital. Hence, the transition parameter from *H* to *A* is *h*_*a*_ = (1 *− p*_4_ *− p*_5_)*/t*_6_.
7. People in ICU (U) may decease (F) or may get better and be transferred to other non-ICU department (HU). These transits are modeled by the linear terms *u*_*f*_ *U* (*t*) and *u*_*hu*_*U* (*t*), respectively. Here we have 2 possible ways for individuals in ICU, die or get better.
  - The transition parameter from *U* to *F* is *u*_*f*_ = *p*_7_*/t*_7_.
  - The parameter that governs the transition from *U* to *HU* is *u*_*hu*_ = (1 *− p*_7_)*/t*_8_.
8. Finally, an individual in *HU* may get better and discharged. This transit is modeled by the linear term *hu*_*a*_*HU* (*t*), where *h*_*ua*_ = 1*/t*_9_.

Since other known coronavirus have a seasonal behaviour [18], as a hypothesis, two main periods have been considered: virulence season (September to April) and non-virulence season (May to August). Due to this, some parameter values will vary depending on the season.

Taking into account the above description of the populations and the model parameters, the following system of difference equations (1) describes the transmission dynamics of COVID-19 in the province of Granada (Spain) over the time.

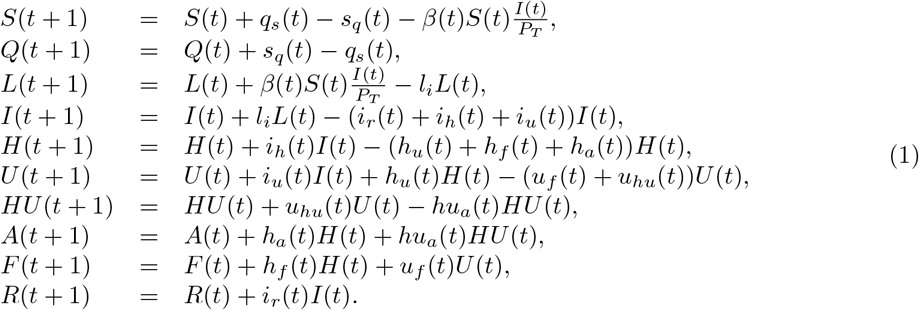

Fig. 1 shows a flow diagram about how individuals may move throughout the different groups with respect to the disease. Note that the right part of Fig. 1 (H, U, F, A, HU) represents the flow of patients within hospital departments. There is not movement between infectious (I) to deceased (F) as there is not data available. Also, we do not consider re-infection.

**Figure 1:**
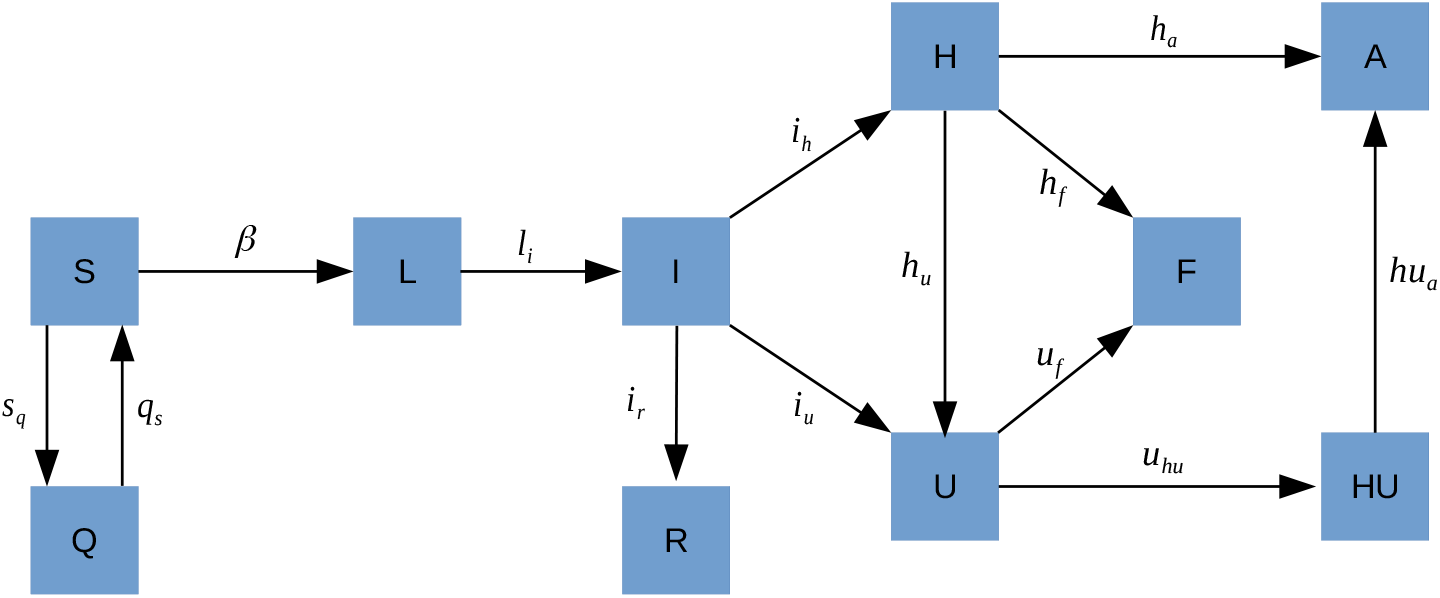
Flow diagram of the COVID-19 transmission dynamics. The boxes represent the groups of individuals respect to the disease, and the letters next to the arrows represent the transition rates between states.

## 3 Model calibration

As we have mentioned before, we gathered data from different hospitals of the Spanish province of Granada with the aim at calibrating the model parameters, in such a way that the model may describe the evolution of the pandemics in this region.

Hence, the model parameters *β, t*_0_, *p*_1_, *t*_1_, *p*_2_, *t*_2_, *p*_4_, *t*_4_, *p*_5_, *t*_5_, *t*_6_, *p*_7_, *t*_7_, *t*_8_ and *t*_9_ have to be calibrated in the intervals described in Table 2. Also, the initial number of latent and infectious to establish an appropriate initial condition the 1 March 2020. According to the data, at 1 March there where 5 people in the hospital circuit and none at ICU or recovered.

**Table 1:** Summary of the known mathematical model parameters.

| Parameter | Transition | Time | Value |
| --- | --- | --- | --- |
| $s_q$ | $S \longrightarrow Q$ | 16 March | 700 000 |
|  |  | 31 March | 150 000 |
| $q_s$ | $Q \longrightarrow S$ | 13 April | 150 000 |
|  |  | 5 May to 21 June | 8 750 |
| $t_3$ | $I \longrightarrow R$ | All simulation | 14 |

**Table 2:** Summary of the mathematical model parameters to be calibrate and their calibration ranges. The calibration ranges for the hospital’s data have been obtained calculating the percentiles 10 and 90 of the data.

| Parameter | Transition | Time | Calibration range |
| --- | --- | --- | --- |
| $I(0)$ | — | 1 March | 100 – 5000 |
| $L(0)$ | — | 1 March | 100 – 5000 |
| $\beta$ | $S \longrightarrow L$ | 1 March to 15 March<br>15 March to 4 May<br>virulence season<br>non-virulence season | 0.07 – 0.7<br>0.03 – 0.4<br>0.03 – 0.4<br>0.03 – 0.4 |
| $t_0$ | $L \longrightarrow I$ | All simulation | 1 – 6 |
| $p_1$ | $I \longrightarrow H$ | virulence season<br>non-virulence season | 0.01 – 0.04<br>0.01 – 0.04 |
| $t_1$ | $I \longrightarrow H$ | All simulation | 9 – 12 |
| $p_2$ | $I \longrightarrow U$ | virulence season<br>non-virulence season | 0.001 – 0.004<br>0.001 – 0.004 |
| $t_2$ | $I \longrightarrow U$ | All simulation | 9 – 12 |
| $p_4$ | $H \longrightarrow U$ | virulence season<br>non-virulence season | 0.000 – 0.139<br>0.000 – 0.119 |
| $t_4$ | $H \longrightarrow U$ | virulence season<br>non-virulence season | 1 – 8.6<br>3.2 – 13.6 |
| $p_5$ | $H \longrightarrow F$ | virulence season<br>non-virulence season | 0.097 – 0.297<br>0.018 – 0.218 |
| $t_5$ | $H \longrightarrow F$ | virulence season<br>non-virulence season | 1 – 15<br>1 – 15.2 |
| $t_6$ | $H \longrightarrow A$ | virulence season<br>non-virulence season | 3 – 15<br>4 – 29 |
| $p_7$ | $U \longrightarrow F$ | virulence season<br>non-virulence season | 0.245 – 0.445<br>*No deceased* |
| $t_7$ | $U \longrightarrow F$ | virulence season<br>non-virulence season | 1 – 23.3<br>*No deceased* |
| $t_8$ | $U \longrightarrow HU$ | virulence season<br>non-virulence season | 6 – 36.2<br>4.7 – 80.1 |
| $t_9$ | $HU \longrightarrow A$ | virulence season<br>non-virulence season | 4 – 25<br>2.7 – 18.9 |

To calibrate the model, we consider four different *β*s: before and after the lockdown, and once the confinement has finished, non-virulence and virulence season behaviour. Also, we take into account that the basic reproduction number of our model *R*_0_ = *β/*(*i*_*r*_ + *i*_*u*_ + *i*_*h*_) lies in reliable values as *t* goes on, time *t* in days.

Our main source data (hospital’s data) collects the number of daily hospitalizations, daily people in ICU, accumulated deaths and accumulated discharges in the hospitals of the province of Granada from 1 March to 22 September 2020. These data allows us to determine intervals where to search the model parameter values during the calibration. Although the mean of the intervals determined by the hospital’s data seems to be the natural option for some of the parameter values, in this case, these mean values may not describe the behavior of the system, since the data do not follow centered distributions. In order to obtain suitable numerical values of the model parameters, the 10 and 90 percentile of the stay time in each state of the hospital circuit (H, U, F, A, HU) are obtained and settled as the calibration range where looking for the proper values of the parameters. The intervals to determine the parameter values, those given by the hospital’s data and the other, can be seen in Table 2 under the column *Calibration range*.

Also, we are going to use the seroprevalence study [13], which determines the percentage of accumulated infected people from the whole population is also available. According to the prevalence study of COVID-19 in Spain, carried out between May and June 2020, the accumulated infected population in the province of Granada from the 27 April to 11 May is in the range [1.7%, 4.4%]. The range from 18 May to 1 June is [2.3%, 4.8%] and the range from 8 June to 22 June is [2.3%, 5.5%].

The model has been calibrated using the optimization algorithm NS for PSO [19] adapted for this problem. Difference between real data and the model outputs - hospitalized at ward, ICU, deceased, discharged and recovered - has been minimized in order to obtain the model parameter values. We have calibrated the model 600 times with 100 000 model evaluations each one, and for all the calibrated sets of model parameter values (all of them valid because lie on the calibration range intervals) we calculate the mean and the 95% confidence interval (CI95%) by the percentiles 2.5 and 97.5 in each time instant *t*. This way, we are able to capture and quantify the possible data uncertainty. The results of the calibration are good enough to represent the effects of the pandemic in Granada during 1 March - 22 September 2020 period (Fig. 2).

**Figure 2:**
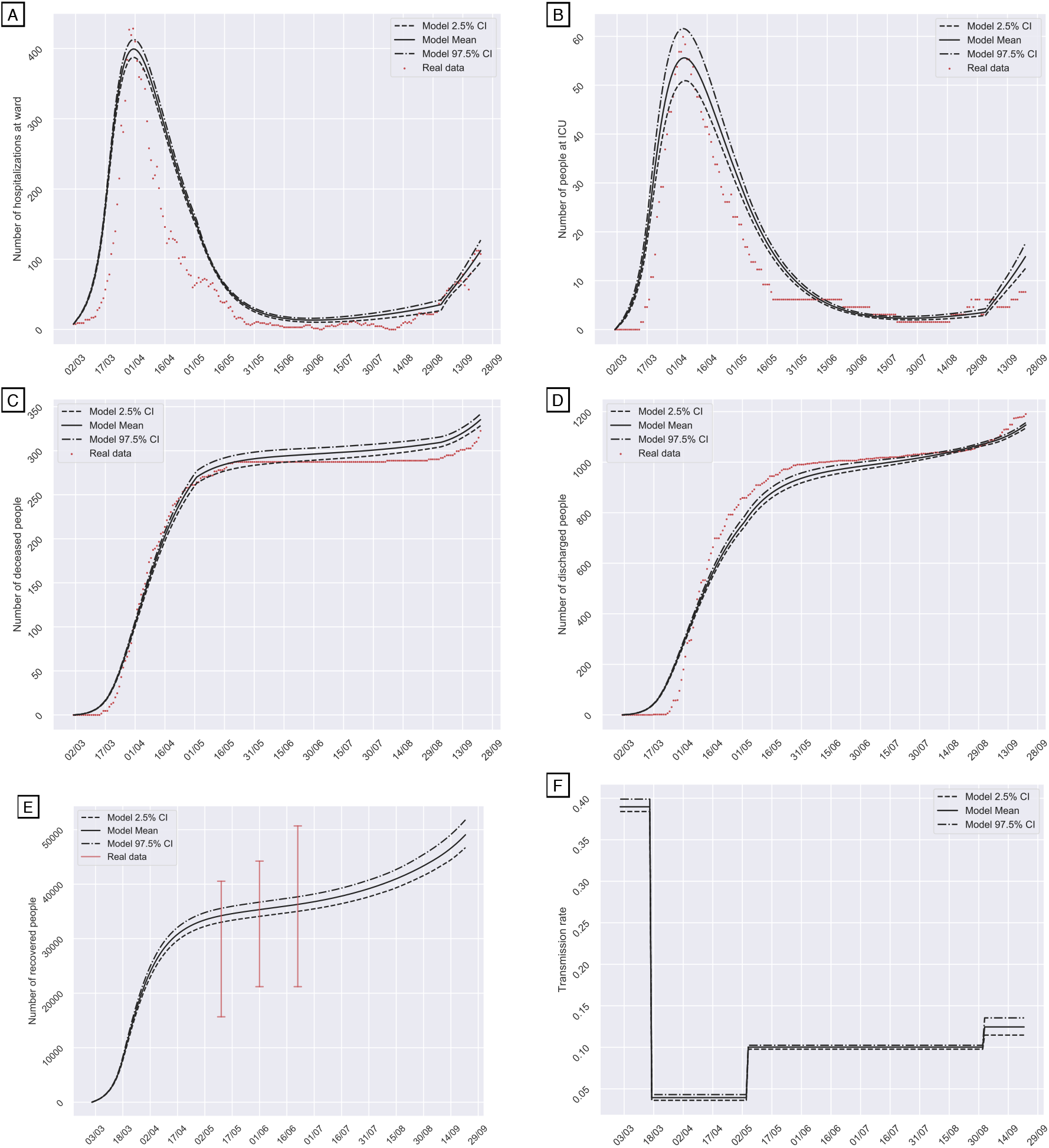
Model calibration. (A) Number of daily hospitalized at ward people. (B) Number of daily people in ICU. (C) Number of accumulated deceased people. (D) Number of accumulated discharged people. (E) Number of accumulated recovered people. (F) Transmission rate - *β*. The black bands are the mean and the 95%CI of the model outputs. The red points are the data gathered from the hospitals. Vertical red lines of (E) show the confidence intervals of the prevalence study [13] carried out in Spain last spring.

### 3.1 Some notes on the calibrated model parameter values

The mean and confidence interval of the calibrated parameters is shown in Table 3. The availability of reliable hospital’s data allowed us to provide realistic intervals to calibrate the model parameters and, once the model has been calibrated, meaningful model parameter values.

**Table 3:** Summary of the calibrated model parameter values.

| Parameter | Transition | Time | Calibrated value<br>Mean (2.5%CI - 97.5%CI) |
| --- | --- | --- | --- |
| $I(0)$ | — | 1 March | 1190 (1165 – 1222) |
| $L(0)$ | — | 1 March | 573 (558 – 590) |
| $\beta$ | $S \rightarrow L$ | 1 March to 15 March | 0.390 (0.383 – 0.399) |
|  |  | 15 March to 4 May | 0.039 (0.036 – 0.043) |
|  |  | virulence season | 0.125 (0.115 – 0.135) |
|  |  | non-virulence season | 0.100 (0.098 – 0.102) |
| $t_0$ | $L \rightarrow I$ | All simulation | 3.822 (3.571 – 3.955) |
| $p_1$ | $I \rightarrow H$ | virulence season | 0.027 (0.026 – 0.028) |
|  |  | non-virulence season | 0.014 (0.010 – 0.019) |
| $t_1$ | $I \rightarrow H$ | All simulation | 11.610 (11.030 – 11.869) |
| $p_2$ | $I \rightarrow U$ | virulence season | 0.00238 (0.00209 – 0.00274) |
|  |  | non-virulence season | 0.00061 (0.00045 – 0.00076) |
| $t_2$ | $I \rightarrow U$ | All simulation | 10.528 (9.446 – 11.528) |
| $p_4$ | $H \rightarrow U$ | virulence season | 0.020 (0.019 – 0.021) |
|  |  | non-virulence season | 0.019 (0.018 – 0.020) |
| $t_4$ | $H \rightarrow U$ | virulence season | 4.333 (4.279 – 4.397) |
|  |  | non-virulence season | 8.309 (8.223 – 8.392) |
| $p_5$ | $H \rightarrow F$ | virulence season | 0.136 (0.135 – 0.138) |
|  |  | non-virulence season | 0.074 (0.071 – 0.082) |
| $t_5$ | $H \rightarrow F$ | virulence season | 8.571 (8.468 – 8.696) |
|  |  | non-virulence season | 7.507 (7.450 – 7.583) |
| $t_6$ | $H \rightarrow A$ | virulence season | 13.525 (13.501 – 13.596) |
|  |  | non-virulence season | 13.244 (13.202 – 13.295) |
| $p_7$ | $U \rightarrow F$ | virulence season | 0.248 (0.254 – 0.243) |
| | | non-virulence season | $4.852 \cdot 10^{-11}$ ($1.202 \cdot 10^{-11}$ – $8.418 \cdot 10^{-11}$) |
| $t_7$ | $U \rightarrow F$ | virulence season | 13.729 (13.611 – 13.798) |
| | | non-virulence season | $2.040 \cdot 10^{+10}$ ($1.140 \cdot 10^{+10}$ – $1.030 \cdot 10^{+11}$) |
| $t_8$ | $U \rightarrow HU$ | virulence season | 14.990 (14.832 – 15.132) |
|  |  | non-virulence season | 19.973 (19.346 – 20.621) |
| $t_9$ | $HU \rightarrow A$ | virulence season | 13.650 (13.603 – 13.697) |
|  |  | non-virulence season | 11.005 (10.924 – 11.081) |

In 1 March 2020 the calibrated number of infectious 1190, *CI*95% 1165 *−* 1222, (0.13%, *CI*95% 0.125% *−*0.133%) and latent 573, *CI*95% 558 *−* 590, (0.062%, *CI*95% 0.060% *−* 0.064%) was low. Data provided by the hospitals in the province of Granada, from 1 March to 15 March, compared with the registry of the previous year did not show an increase of respiratory disease cases that could be attributed to COVID-19, what it is in accordance with our estimation of the low number of COVID-19 patients at the beginning of the pandemic (1 March 2020). Later, the number of infected patients experienced a great increase, what supports that the effective reproduction number *R*_0_ was initially very high, (5.42, *CI*95% 5.35 *−* 5.54), value which is in accordance with Sanche et al [20].

After the beginning of the lockdown declared by the Spanish government in 14 March, the number of infected patients kept growing as a consequence of the contagions produced in the previous weeks with high *β*. Nevertheless, the basic reproductive number *R*_0_ decreased to (0.55 CI95% 0.504 *−* 0.597), not only because of lockdown but also the social distancing, the use of facemasks and other population measures, reducing the *β* (lower right graphic of Fig. 2).

During the non-virulence season after the lockdown is finished, the incidence of the disease is kept low, with a stabilization of the number of hospitalizations and deceases. The *R*_0_ value is low, (1.40 CI95% 1.367*−* 1.427), but it increases until slightly higher values than at the beginning of the virulence season (*R*_0_ = 1.73, CI95% 1.597*−* 1.879). This can be observed with the increment of hospitalized and intensive care unit people, together with the end of the stagnation in the number of deceased and discharged on Figure 2.

Different values of the parameters can be appreciated, more precisely the percentage of infected people who needs hospitalization (hospitalization at ward *p*_1_, or ICU treatment *p*_2_). During the virulence season, about 2% of the infected people need hospitalization at ward, and 0.2% of the infected people enter the ICU directly without passing through the hospitalization ward. During the non-virulence season, about 1.4% of the infected people need hospitalization at ward, and 0.06% of the infected people enter the ICU directly without passing through the hospitalization ward.

## 4 Vaccination

Once the model has been calibrated, we can simulate different possible scenarios for the future. In particular, taking into account that several vaccines candidates are being created, we are going to deal with vaccination strategies. In order to simulate the effect of the vaccine in the population of the Spanish region of Granada, a new state has to be considered in the mathematical model:

- **(V)**vaccinated, when the individual is vaccinated and the vaccine is effective, protecting the individual.

We suppose that only susceptible people are vaccinated. The vaccine protects against the most dangerous symptoms of the COVID-19, and stops the spread of the disease. However, the effect of the vaccine is not permanent and the vaccinated people may become susceptible again. The transit from *S* to *V* is modeled by the term *s*_*v*_(*t*) and the transit from *V* to *S*, it is modeled by the term *v*_*s*_(*t*). Both parameters describe the number of people who are moving from one state to other, at time *t*.

The mathematical system of difference equations, which includes the vaccination of the population is shown in system (2). In black, the new terms.

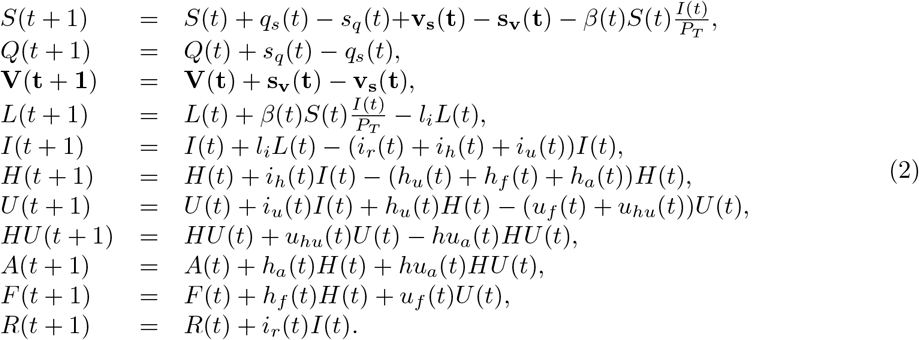

The administration capacity of vaccination is limited and it is not possible to vaccinate the whole population on the same day. We assume that, being this pandemic an extraordinary situation, it would be possible to vaccinate 1% of the population every day.

Not every vaccinated person acquires protection against the disease. We consider the vaccine effectiveness as the percentage of vaccinated people who acquire immunity. Also, the duration effect of the vaccine is unknown, and we will assume an immunity of a year on all cases.

To simulate the possible scenarios for the population vaccination, as it is still not known the possible effectiveness of the future COVID-19 vaccines, 100%, 75%, 50% and 25% of effectiveness have been compared with the non vaccination scenario. All the scenarios begin to administer the vaccine on 1 December 2020. The results of this simulation have been presented in Figure 3.

**Figure 3:**
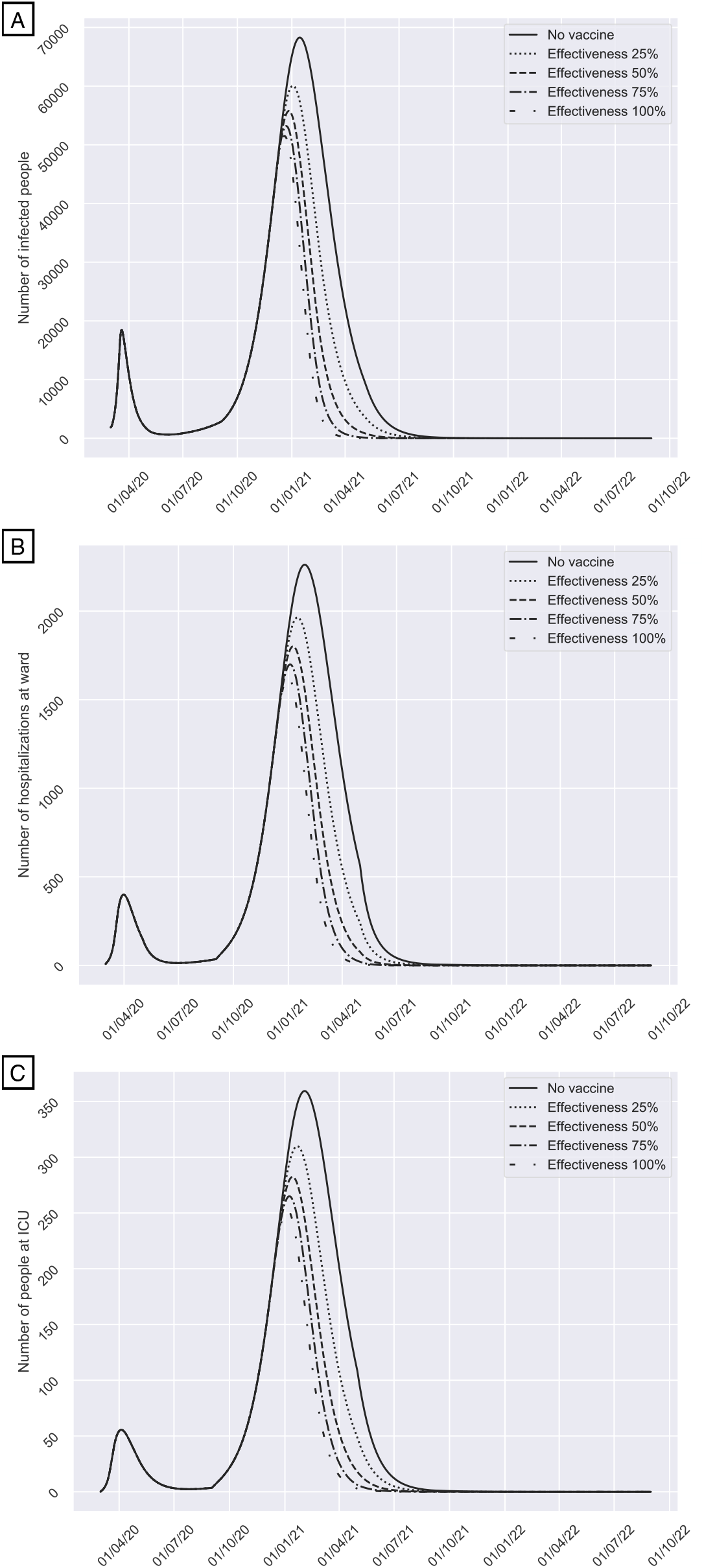
COVID-19 evolution depending on the vaccination effectiveness (starting on 1/12/2020): No vaccination, 25%, 50%, 75% 100%. (A) Number of infected people. (B) Number of daily hospitalized at ward people. (C) Number of daily people in ICU. The percentage of infected population on the peak is 7.4%, 6.5%, 6.1%, 5.7%, 5.6% for no vaccination and a vaccine effectiveness of 25%, 50%, 75% 100%, respectively.

On Figure 3, the difference of the vaccine effectiveness effect is shown. As the effectiveness of the vaccine increases, the height of the peak decreases. It is important to remark the importance of achieving 50% of vaccine effectiveness, since the major reduction is seen from that point. The difference between the hospitalization peak from the scenario of non vaccination and the scenario of 50% effectiveness is 20.6% of reduction, meanwhile the difference on the hospitalization peak from the scenario of 50% effectiveness to the scenario of 100% effectiveness is only 9.6% of reduction. This results are in accordance with the WHO recommendation [21].

The availability of the vaccine is still unknown, but, according to some declarations of the Spanish government [22], it could be available at the beginning of December 2020. Anyway, we have simulated two availability scenarios: December 2020 and February 2021, assuming a vaccine effectiveness of 50%. The election of the vaccine effectiveness is based on the WHO recommendation where it is considered a substantial progress a vaccine effectiveness of 50% [21]. The results have been compared with no vaccination scenario in Figure 4.

**Figure 4:**
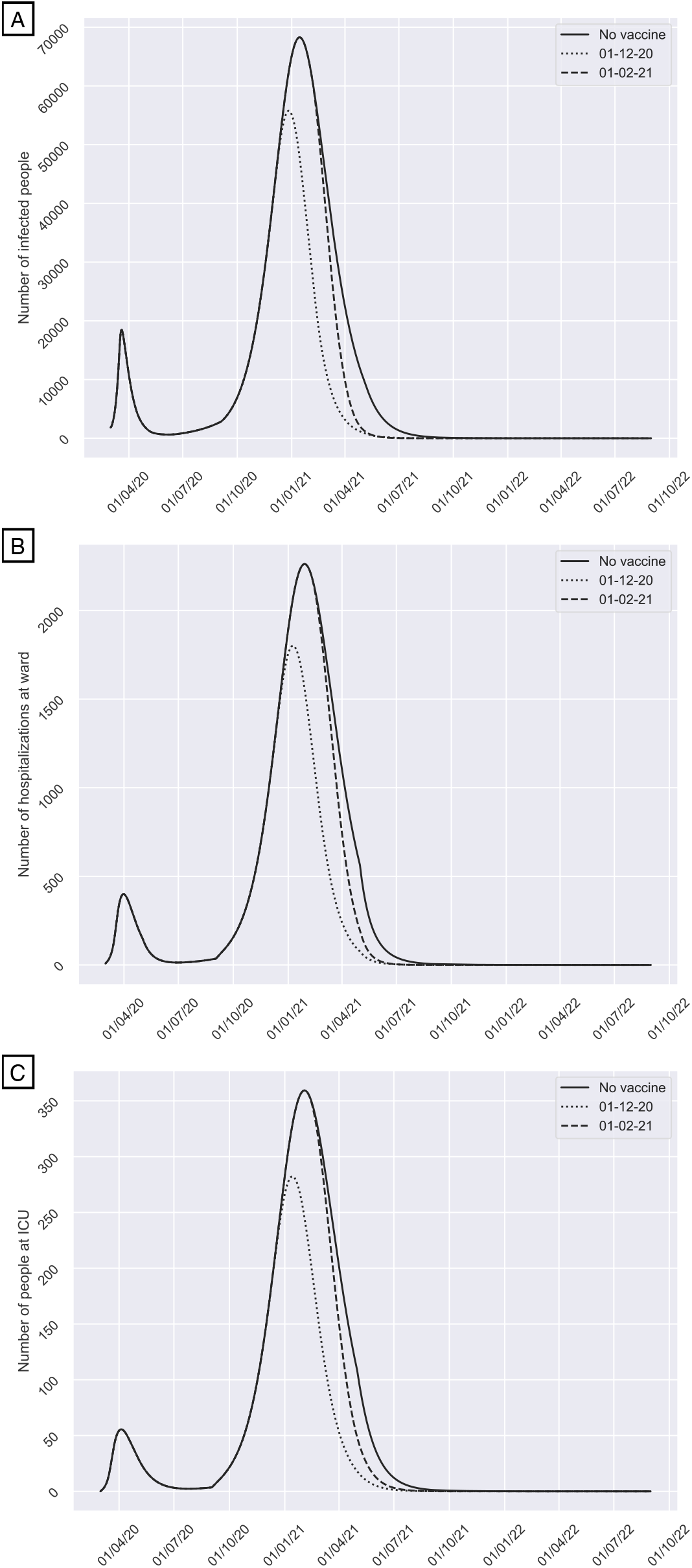
COVID-19 evolution depending on the vaccination start date (with a 50% of effectiveness): No vaccination, vaccination from 1/12/2020 and vaccination from 1/2/2021. (A) Number of infected people. (B) Number of daily hospitalized at ward people. (C) Number of daily people in ICU. The difference between starting the vaccination campaign on 1 December 2020 or on 1 February 2021 is remarkable.

As it can be seen in Figure 4, the difference between starting the vaccination campaign on 1 December 2020 or on 1 February 2021 is quite remarkable. There is 18.3% of reduction of infected people, 20.4% of reduction in hospitalized and 21.4% of reduction in ICU on the pandemics’ peak. There is also a time advance of 19 days on the infected and hospitalized peak and a time advance of 20 days on the ICU peak if the vaccination begins in December 1st instead of February 1st. It should be also noticed that, if the vaccination starts on 1 February 2021, the difference between the vaccination and non vaccination scenarios is only seen at the end of the pandemics, with no affectation on the peak.

It is important to remark that, according the the date of release of the vaccine, the effect on this winter pandemic will be reduced. Herd immunity effect will be limited since the infection evolves faster than the vaccination campaign. Because of this fact, it would be necessary to increase and combine health measures in order to decrease the spread of the illness until a vaccine could be available to the population.

## 5 Discussion

Coronaviruses cause respiratory and intestinal diseases in many animal species. In humans, cold-like symptoms and upper respiratory tract infections can be caused by 4 existing coronaviruses, including 2 alpha coronaviruses (229E and NL63) and 2 beta coronaviruses (OC43 and HKU1). In addition, there are 3 beta coronaviruses that cause severe respiratory syndromes in humans (SARS-CoV-1, MERS-CoV, and SARS-CoV-2) [23]. Main treatment options suggested against SARS-CoV-2 include chloroquine/hydroxychloroquine, favipiravir, remdesivir, lopinavir/ritonavir, umifenovir, steroids, cepharanthine, interferon, ivermectin, ribavirin, convalescent plasma, anticoagulants, mesenchymal stem cells and neutralizing antibodies, among others. However, there is an urgent need for effective rapid diagnostics, vaccines and therapeutics to detect, prevent, treat and contain SARS-CoV-2 [24, 25, 26].

Explanatory mathematical models of diseases must allow to predict the evolution of the disease considering a wide range of possible scenarios. It must be able to estimate the healthcare demands that will have to be covered by hospitals early enough as to implement management and operative adaptations regarding multiple aspects, including the extension of ICU to other places that could be equipped and be used for the management of the COVID-19 patients, as well as the reservation of out-of-hospital stays, such as hotels, for health personnel or even for patients with low severity who must be lockdown in a controlled context [27].

In this paper we propose a mathematical model to study the current transmission dynamics of SARS-CoV-2. It is a SEIR model where the circuit of patients moving inside the hospital dependencies, the particular features of the disease and the asymptomatic infectious, a principal source of contagiousness [28], are considered, returning a more precise portrait of the use of hospital resources. We tested the model using registered cases and population data from the hospitals of the Spanish province of Granada that represent a size of near 1 million citizens. In this province, there are low density of population and low prevalence of COVID-19 on the spring 2020 outbreak.

A direct and permanent line of communication is important for the transmission of data in real time between the heads of hospital centres and the teams in charge of analysing the epidemiological evolution of the disease, especially through tools such as the one presented in this paper. This will ensure the optimization of future predictions on the basis of the augmentation of the knowledge database, contributing to face the fluctuations that may appear in the transmission of the virus depending on variables that are not sufficiently clarified at the moment.

One of the strong aspects of our model is that it has been possible to calibrate and validate from data that has been collected and reported through an informative protocol agreed at the province of Granada, which ensures homogeneity as well as prevents biases that may appear on the count of each type of subject, consideration of infected by diagnostic, etc; also, the model can be fed with new data to update the predictions regularly.

Another important aspect is that our model is able to distinguish patients recovered from COVID-19 and those who have died from the disease, since it will depend on the behaviour of the virus in the future, so that, the group of recovered can be considered as a group immune to infection, or they may again be considered as a susceptible population in the event of significant phenotypic changes in the virus or possible reinfections.

Our simulations show that we have to keep population measures in order to control the pandemic and avoid a new concerning outbreak. After the beginning of the lockdown declared by the Spanish government in 14 March, the number of infected patients kept growing as a consequence of the contagions produced in the previous weeks of high *β*. However, the outbreak could be delayed if the disease *β* is reduced as much as possible, eluding the coincidence with the influenza’s seasonal peak. This will permit a better planning of the hospital resources, preparing a COVID-19 area without reducing the care attention to patients with other diseases, preventing the collapse of the health services, as happened last March-April 2020.

The transmission capacity of the virus at the beginning of the pandemic was very high, with a *β* close to 0.4 and a *R*_0_ greater than 5. Numerous studies have stated the effectiveness of non-pharmaceutical measures in reducing the transmission of respiratory virus [29, 30, 31, 32, 33, 34, 35, 36, 37, 38]. Then, *β* and *R*_0_ decreased significantly, reaching an average of 1.40 and making it possible to control the pandemic by approaching to 1. The eradication of the virus in our environment is not to be expected in the short term, since it has not been possible to implement socio-sanitary measures that reduce the *R*_0_ and its confidence interval of potential values below 1.

By 2 October 2020, there are 42 candidate vaccines for SARS-CoV-2 in clinical evaluation and 151 in preclinical models [21] that have been developed using a wide range of technologies and strategies, such as RNA, DNA, nanoparticles, recombinant proteins, live attenuated vaccines, inactivated vaccines, synthetic and modified virus-like particles [25, 39]. However, the impact of the COVID-19 vaccines on infection, re-infection, transmission, and disease severity has not been presented yet as conclusions of clinical trials [40]. Computational model of SARS-CoV-2 and vaccination in USA revealed that to extinguish the ongoing epidemic without other non-pharmaceutical interventions, the vaccine efficacy has to achieve at least 60% when coverage is 100%, and at least 80% when coverage is up to 75% to drop the peak by 85%-86%, 61%-62%, and 32% depending on the population previously exposed to the virus (5%, 15%, and 30%, respectively) [41]. WHO recommends a vaccine efficacy greater than 30% in 95% confidence interval estimation, and considers that an efficacy around 50% would represent substantial progress [40, 42].

The evolution of the pandemic with the current number of infected people and transmission rate, even though if an effective vaccine would be available, seems to be rising. More health policies must be implemented in order to decrease the transmission rate until the vaccine will be available for the whole population. Moreover, more vulnerable population groups (such as old people) should be vaccinated as soon as possible, since herd effect might be difficult to obtain before an outbreak, unless more drastic health policies are implemented.

Finally, we have to take into account that, as shown in Figure 4, in summer 2021 when we reach low levels of infections, the social distancing, facemasks and other non-pharmaceutical measures cannot be removed suddenly. Otherwise, the transmission rate increases, also *R*_0_, and a new outbreak may arise (as it can be seen in Figure 5) at the beginning of 2022.

**Figure 5:**
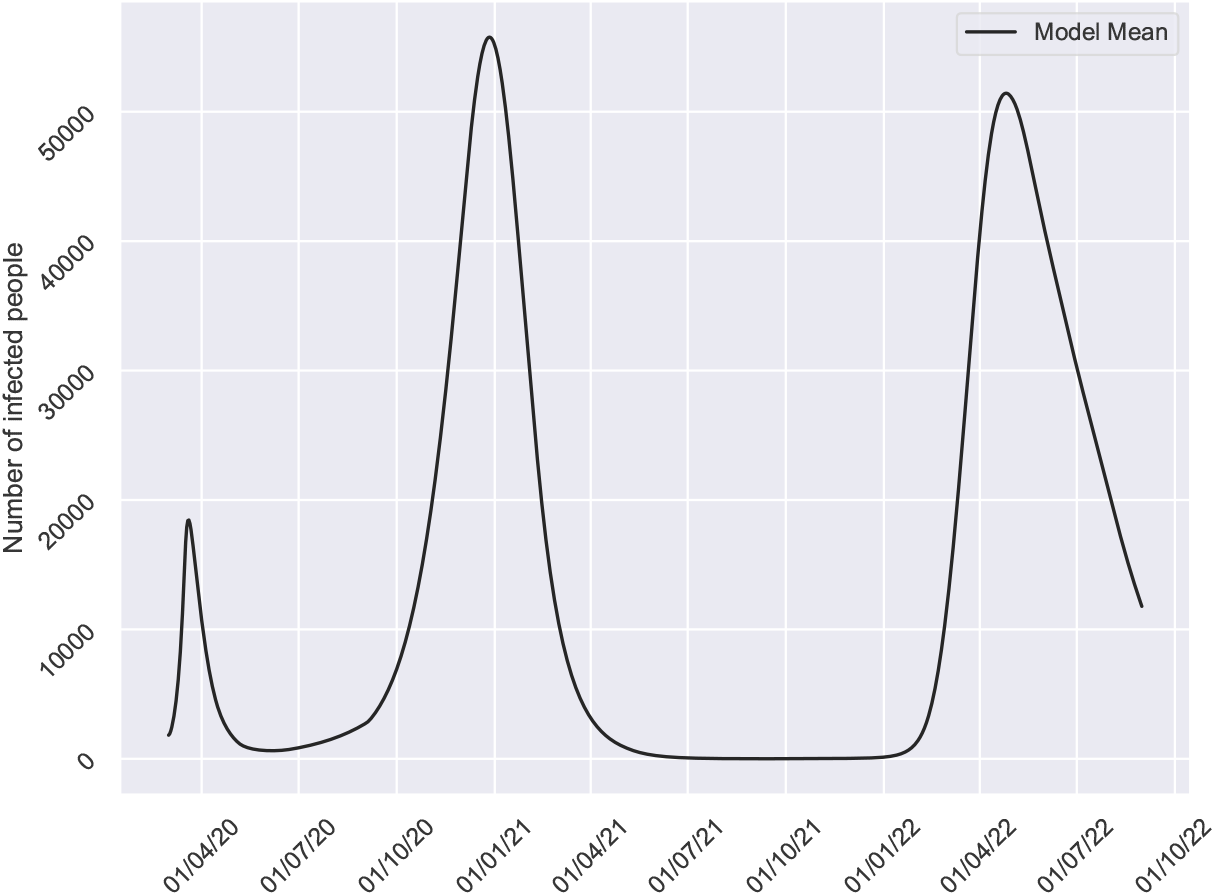
COVID-19 evolution with the initial transmission rate from September 2021 due to the sudden end of non-pharmaceutical measures. Vaccine campaign 2020 starting on 1 December 2020 with 50% effectiveness and a year effect and no vaccination campaign in 2021.

## 6 Conclusion

The modified SEIR model presented in this paper can be a useful tool for providing insight into the transmission dynamics of SARS-CoV-2. It considers the circuit of patients moving inside the hospital dependencies, returning a more precise portrait of the use of hospital resources, in a region with low density of population and low COVID-19 prevalence. Once the model is calibrated, vaccination simulations have been performed to evaluate the evolution of COVID-19. The simulations show that the number of infected is going to increase reaching a peak in mid December - mid January. Also, the vaccination should be combined with other health measures to face COVID-19 and avoid overloading in the health system. Furthermore, our model supports the WHO recommendation about the effectiveness of the vaccine, that should be, at least, of 50%. Moreover, when the next outbreak declines, we will not yet be out of danger, and we will have to resume life as we knew it before the pandemic relaxing measures of social situation and masks in a gradual and controlled way to avoid a new outbreak.

The predictions may provide enough time to the health systems to establish the appropriate measures to face outbreaks that could collapse human and material resources. Moreover, future pharmaceutical and non-pharmaceutical interventions, as well as aspects related to SARS-CoV-2 biology and behaviour, can be represented through the model parameters to act as modulator of the *β* what provide the required flexibility to adapt the model throughout the time.

### Limitations of the model

The proposed model is a classical system of difference equations. For its building, we assume usual hypothesis as the homogeneous mixing of the population (any individual may infect any individual). No age groups are considered, and the hospitalization and decease rate may vary depending on the age of the individuals. Also, we assume that all lockdown people are susceptible, when latent, asymptomatic infectious and recovered are also in lockdown. Furthermore, mobility and other spatial aspects are not considered.

Moreover, it is important to consider that the model must be particularized for each population, either provincial or larger, since the transmission rate must respond to the effect of health policy measures and the response of the population to the recommendations, such as opening educational centers, social distancing, use of face masks, adaptation of public and private services to establish physical barriers that hinder the spread of the virus, etc.

## Supporting information

Ethics committee approval

## Data Availability

All data used in the study is provided in the manuscript.

