## Supplementary material for "COVID-19 in regions with low prevalence and low density of population. An uncertainty dynamic modeling approach": Ethics committee approval

### DICTAMEN ÚNICO EN LA COMUNIDAD AUTÓNOMA DE ANDALUCÍA

D/Dª: Francisco Manuel Luque Martínez como secretario/a del CEIM/CEI Provincial de Granada

#### CERTIFICA

Que este Comité ha evaluado la propuesta del promotor/investigador (No hay promotor/a asociado/a) para realizar el estudio de investigación titulado:

| | |
|---|---|
| TÍTULO DEL ESTUDIO: | MODELIZACIÓN MATEMÁTICA DINÁMICA PARA EL SEGUIMIENTO DE LA EVOLUCIÓN DE LA COVID-19 EN LA PROVINCIA DE GRANADA ,( COVID-19 Granada) |
| Protocolo, Versión: | 1 |
| HIP, Versión: | 1 |
| CI, Versión: | |

Y que considera que:

Se cumplen los requisitos necesarios de idoneidad del protocolo en relación con los objetivos del estudio y se ajusta a los principios éticos aplicables a este tipo de estudios.

La capacidad del/de la investigador/a y los medios disponibles son apropiados para llevar a cabo el estudio.

Están justificados los riesgos y molestias previsibles para los participantes.

Que los aspectos económicos involucrados en el proyecto, no interfieren con respecto a los postulados éticos.

Y que este Comité considera, que dicho estudio puede ser realizado en los Centros de la Comunidad Autónoma de Andalucía que se relacionan, para lo cual corresponde a la Dirección del Centro correspondiente determinar si la capacidad y los medios disponibles son apropiados para llevar a cabo el estudio.

Lo que firmo en Granada a 21/10/2020

D/Dª. Francisco Manuel Luque Martínez, como Secretario/a del CEIM/CEI Provincial de Granada

### CERTIFICA

Que este Comité ha ponderado y evaluado en sesión celebrada el 26/10/2020 y recogida en acta 09/20 la propuesta del/de la Promotor/a (No hay promotor/a asociado/a), para realizar el estudio de investigación titulado:

| | |
|---|---|
| TÍTULO DEL ESTUDIO: | MODELIZACIÓN MATEMÁTICA DINÁMICA PARA EL SEGUIMIENTO DE LA EVOLUCIÓN DE LA COVID-19 EN LA PROVINCIA DE GRANADA ,( COVID-19 Granada) |
| Protocolo, Versión: | 1 |
| HIP, Versión: | 1 |
| CI, Versión: | |

Que a dicha sesión asistieron los siguientes integrantes del Comité:

**Presidente/a**

D/Dª. José Dario Sánchez López

**Vicepresidente/a**

D/Dª.

**Secretario/a**

D/Dª. Francisco Manuel Luque Martínez

**Vocales**

D/Dª. Jesús Martínez Tapias
D/Dª. Juan Ramón Delgado Pérez
D/Dª. Berta Gorlat Sánchez
D/Dª. José Cabeza Barrera
D/Dª. Sonia Dominguez Almendros
D/Dª. Juan Mozas Moreno
D/Dª. José Uberos Fernández
D/Dª. MARIA ESPERANZA DEL POZO GAVILAN
D/Dª. AURORA BUENO CAVANILLAS
D/Dª. Paloma Muñoz de Rueda
D/Dª. Esther Espínola García
D/Dª. ANTONIO MORALES ROMERO
D/Dª. Encarnación Martínez García
D/Dª. FRANCISCO LUIS MANZANO MANZANO
D/Dª. MIGUEL LÓPEZ GUADALUPE
D/Dª. JUAN ROMERO COTELO
D/Dª. MANUEL MARTIN DIAZ
D/Dª. JOSÉ LUIS MARTÍN RODRÍGUEZ
D/Dª. Juan Díaz García
D/Dª. LUIS MIGUEL DOMENECH GIL
D/Dª. Luis Javier Martínez González
D/Dª. JESÚS CARDONA CONTRERAS
D/Dª. Pilar Guijosa Campos
D/Dª. José Luis Martín Ruiz
D/Dª. MARÍA DOLORES GARCÍA VALVERDE
D/Dª. ESTHER MOLINA RIVAS
D/Dª. ANTONIO JUAN PÉREZ FERNÁNDEZ
D/Dª. MARIANA FÁTIMA FERNÁNDEZ CABRERA
D/Dª. JOAQUINA MARTINEZ GALAN

Que dicho Comité, está constituido y actua de acuerdo con la normativa vigente y las directrices de la Conferencia Internacional de Buena Práctica Clínica.

Lo que firmo en Granada a 21/10/2020
